# Genomic epidemiology of SARS-CoV-2 spread in Scotland highlights the role of European travel in COVID-19 emergence

**DOI:** 10.1101/2020.06.08.20124834

**Authors:** Ana da Silva Filipe, James Shepherd, Thomas Williams, Joseph Hughes, Elihu Aranday-Cortes, Patawee Asamaphan, Carlos Balcazar, Kirstyn Brunker, Stephen Carmichael, Rebecca Dewar, Michael D. Gallagher, Rory Gunson, Antonia Ho, Natasha Jesudason, Natasha Johnson, E Carol McWilliam Leitch, Kathy Li, Alasdair MacLean, Daniel Mair, Sarah E. McDonald, Martin McHugh, Jenna Nichols, Marc Niebel, Kyriaki Nomikou, Richard J Orton, Áine O’Toole, Massimo Palmarini, Yasmin A. Parr, Andrew Rambaut, Stefan Rooke, Sharif Shaaban, Rajiv Shah, Joshua B. Singer, Katherine Smollett, Igor Starinskij, Lily Tong, Sreenu Vattipally, Elizabeth Wastnedge, David L Robertson, Matthew T.G. Holden, Kate Templeton, Emma C. Thomson, on behalf of the COG-UK consortium

## Abstract

SARS-CoV-2, the causative agent of COVID-19, emerged in Wuhan, China in December 2019 and spread rapidly throughout the world. Understanding the introductions of this new coronavirus in different settings may assist control efforts and the establishment of frameworks to support rapid response in future infectious disease outbreaks.

We investigated the first four weeks of emergence of the SARS-CoV-2 virus in Scotland after the first case reported on the 1st March 2020. We obtained full genome sequences from 452 individuals with a laboratory-confirmed diagnosis of COVID-19, representing 20% of all cases until 1st April 2020 (n=2310). This permitted a genomic epidemiology approach to study the introductions and spread of the SARS-2 virus in Scotland.

From combined phylogenetic and epidemiological analysis, we estimated at least 113 introductions of SARS-CoV-2 into Scotland during this period. Clusters containing multiple sequences suggestive of onward transmission occurred in 48/86 (56%). 42/86 (51%) clusters had no known international travel history indicating undetected introductions.

The majority of viral sequences were most closely related to those circulating in other European countries, including Italy, Austria and Spain. Travel-associated introductions of SARS-CoV-2 into Scotland predated travel restrictions in the UK and other European countries. The first local transmission occurred three days after the first case. A shift from travel-associated to sustained community transmission was apparent after only 11 days. Undetected introductions occurred prior to the first known case of COVID-19. Earlier travel restrictions and quarantine measures might have resulted in fewer introductions into Scotland, thereby reducing the number of cases and the subsequent burden on health services. The high number of introductions and transmission rates were likely to have impacted on national contact tracing efforts. Our results also demonstrate that local real-time genomic epidemiology can be used to monitor transmission clusters and facilitate control efforts to restrict the spread of COVID-19.

**Funding:** MRC (MC UU 1201412), UKRI/Wellcome (COG-UK), Wellcome Trust Collaborator Award (206298/Z/17/Z – ARTIC Network; TCW Wellcome Trust Award 204802/Z/16/Z

**Research in context:** *Evidence before this study:* Coronavirus disease-2019 (COVID-19) was first diagnosed in Scotland on the 1^st^ of March 2020 following the emergence of the causative severe acute respiratory system coronavirus 2 (SARS-CoV-2) virus in China in December 2019. During the first month of the outbreak in Scotland, 2310 positive cases of COVID-19 were detected, associated with 1832 hospital admissions, 207 intensive care admissions and 126 deaths. The number of introductions into Scotland and the source of those introductions was not known prior to this study.

*Added value of this study:* Using a combined phylogenetic and epidemiological approach following real-time next generation sequencing of 452 SARS-CoV-2 samples, it was estimated that the virus was introduced to Scotland on at least 113 occasions, mostly from other European countries, including Italy, Austria and Spain. Localised outbreaks occurred in the community across multiple Scottish health boards, within healthcare facilities and an international conference and community transmission was established rapidly, before local and international lockdown measures were introduced.

## Background

The pandemic virus severe acute respiratory system coronavirus 2 (SARS-CoV-2) has spread rapidly throughout the world, including Europe, following its emergence in Wuhan, China in December 2019 (1-4). SARS-CoV-2 is a highly transmissible *Betacoronavirus*, related to the first SARS virus (5). It causes the clinical syndrome coronavirus disease-2019 (COVID-19), characterised by nonspecific respiratory or gastrointestinal viral symptoms and anosmia. In severe cases, acute respiratory distress syndrome, cardiovascular disease, neurological manifestations, thrombosis and renal failure may occur (6, 7). A rare Kawasaki-like disease has recently been described in children (8). Despite the mobilisation of significant resources to contain the outbreak, COVID-19 was declared a Public Health Emergency of International Concern (PHEIC) by the World Health Organisation on 30th January 2020 and a pandemic on 11th March 2020 (9, 10). Many countries are now responding to large outbreaks triggering unprecedented social and economic disruption and challenges to local healthcare systems.

The WHO has declared a PHEIC on five occasions since 2009, all as a result of RNA virus outbreaks (H1N1 influenza, Zika, polio and Ebola). Pathogen genomic sequencing is now established as a core component of the modern epidemiological response to such outbreaks, driven by modern nucleic acid sequencing technologies that can immediately yield entire pathogen genomes from clinical samples (11). The integration of viral genomic data with spatial, temporal and other metadata in a genomic epidemiology framework has allowed enhanced inference of the origin and transmission dynamics of disease outbreaks (12-14). Such an approach is particularly applicable to RNA viruses, as their relatively low-fidelity replication cycle generates mutations in the viral genome at a rate observable over the rapid time scale of an outbreak (15).

In this study, we sequenced laboratory-confirmed positive cases of COVID-19 in Scotland, and analysed them alongside available international data, in order to estimate the number of introduction events and early spread of SARS-CoV-2 in the country. We also aimed to investigate outbreaks in health-care settings in real-time. During the sampling period, 2310 positive cases of COVID-19 were detected (https://statistics.gov.scot), associated with 1832 hospital admissions, 207 intensive care admissions and 126 deaths. Applying a genomic epidemiology approach to the data, we demonstrate the outbreak is the result of multiple separate introductions of the virus associated with international travel, and that community transmission was quickly established in Scotland, well before the introduction of ‘lockdown’ countermeasures on the 23^rd^ March.

## Methods

### Samples

Up to 300 samples per week were selected prospectively following ethical approval from the relevant national biorepository authorities (16/WS/0207NHS and10/S1402/33) between 1^st^ March and 1st April 2020. 50% of samples were randomly selected to achieve a representative target for all Scottish health boards and 50% to cover suspected healthcare-related nosocomial infections as they occurred. Health boards with a small population size were reported at a minimum of 5 sequences per region to avoid deductive disclosure. The Royal Infirmary of Edinburgh and West of Scotland Specialist Virology Centre, NHSGGC conducted diagnostic real-time RT-PCR to detect SARS-CoV-2 positive samples, following nucleic acid extraction utilising the NucliSENS® EasyMag® and Roche MG96 platforms(16). Residual nucleic acid from 452 samples underwent whole genome next generation sequencing at the MRC-University of Glasgow Centre for Virus Research (CVR) and the Royal Infirmary of Edinburgh. Clinical details including recent travel history were obtained from assay request forms submitted to the diagnostic laboratory and where available electronic patient records.

### Sequencing: Oxford Nanopore Technologies (ONT)

Following extraction, samples underwent DNase treatment (AM2222) and libraries were prepared utilising protocols developed by the ARTIC network (v1 and v2) https://artic.network/ncov-2019. 50 fmol of library pools were loaded onto each flow cell (R9.4.1). Sequencing was conducted in MinKNOW version 19.12.5. Raw FAST5 files were basecalled using Guppy version 3.4.5 in high accuracy mode using a minimum quality score of 7. RAMPART v1.0.5 was used to visualise read mapping in real-time. Reads were size filtered, demultiplexed and trimmed with Porechop (https://github.com/rrwick/Porechop), and mapped against reference strain Wuhan-Hu-1 (MN908947). Variants were called using Nanopolish 0.11.3 and accepted if they had a log-likelihood score of greater than 200 and minimum read coverage of 20.

### Sequencing: Illumina MiSeq

Amplicons were generated as described above. DNA fragments were cleaned using AMPURE beads (Beckman Coulter) and 40 ng used to prepare Illumina sequencing libraries with a DNA KAPA library preparation kit (Roche). Indexing was carried out with NEBNext multiplex oligos (NEB), using 7 cycles of PCR. Libraries were pooled in equimolar amounts and loaded on a MiSeqV2 cartridge (500 cycles). To estimate sequencing error rate, seven samples were processed in duplicate from extracted RNA with a target enrichment protocol using NimbleGen probes (Roche). Reads were trimmed with trim_galore (http://www.bioinformatics.babraham.ac.uk/projects/trim_galore/) and mapped with BWA (17) to the Wuhan-Hu-1 (MN908947) reference sequence, followed by primer trimming and consensus calling with iVar (18) and a minimum read coverage of 10.

### Sequence Data

Consensus sequences with >90% coverage were included. All consensus genomes are available from the GISAID database (https://www.gisaid.org), the COG-UK consortium website (https://www.cogconsortium.uk/data/) and BAM files from the European Nucleotide Archive’s [Sequence Read Archive](https://www.ebi.ac.uk/ena/browser/home) service, BioProject PRJEB37886 (https://www.ebi.ac.uk/ena/data/view/PRJEB37886). See supplementary table 1 for IDs and dates of sampling.

### Phylogenetic analysis

Sequences from GISAID were downloaded on the 10^th^ April and aligned in CoV-GLUE (http://cov-glue.cvr.gla.ac.uk). Consensus sequences were aligned using MAFFT (19). A maximum-likelihood phylogenetic tree was constructed using IQ-TREE with the HKY nucleotide substitution model as determined by modeltest-NG (20, 21) and visualised with Figtree. PANGOLIN (https://github.com/hCoV-2019/pangolin) was used for assigning phylogenetic lineages with the additional step of removal of identical sequences (22). The estimated number of introductions into Scotland was calculated by combining lineage assignment with travel history and date of sampling to allow for multiple travel-associated introductions with identical sequences. A Scottish-specific cluster analysis was carried out using the following rules: identical sequences from individuals residing in Scotland were assumed to be linked unless there was a history of travel outside Scotland. Scottish lineages were identified where a basal Scottish sequence was present. Clusters including at least two Scottish sequences were included as evidence of onward transmission if no travel history was present for at least one individual.

### Findings

### Multiple introductions of SARS-CoV-2 in Scotland

452 SARS-CoV-2 genomes were generated with >90% genome coverage, representing 20% of laboratory-confirmed Scottish COVID-19 cases. The median Ct value was 28.98 (range 16-39). Of 452 individuals, 272 (60%) reported no travel, and 61 (14%) reported travel outside Scotland. No travel history was recorded for 119 (26%) individuals. Countries visited included Italy (n=37), Spain (n=7), Austria (n=5), Switzerland (n=3), France (n=2), England (n=1), Ireland (n=1), Poland (n=1) and Thailand (n=1). Three individuals returned from Caribbean cruise holidays.

The first case of COVID-19 in Scotland was a 51-year-old male from Tayside with mild respiratory symptoms who was tested on the 28^th^ February 2020 (and reported positive on the 1^st^ March 2020). He had returned from Italy after attending a rugby match 9 days earlier (23). The first confirmed case who had not travelled occurred three days later, on 2^nd^ March 2020. Reflecting the change from returning travellers to an older disease-susceptible demographic, the median age of cases increased from 44 (IQR 32-51) in the first week of the epidemic to 62 (IQR 47-76) in the fourth, as infections moved from travel-associated to local community transmission (Figure 1; p<0.001).

**Figure 1.**
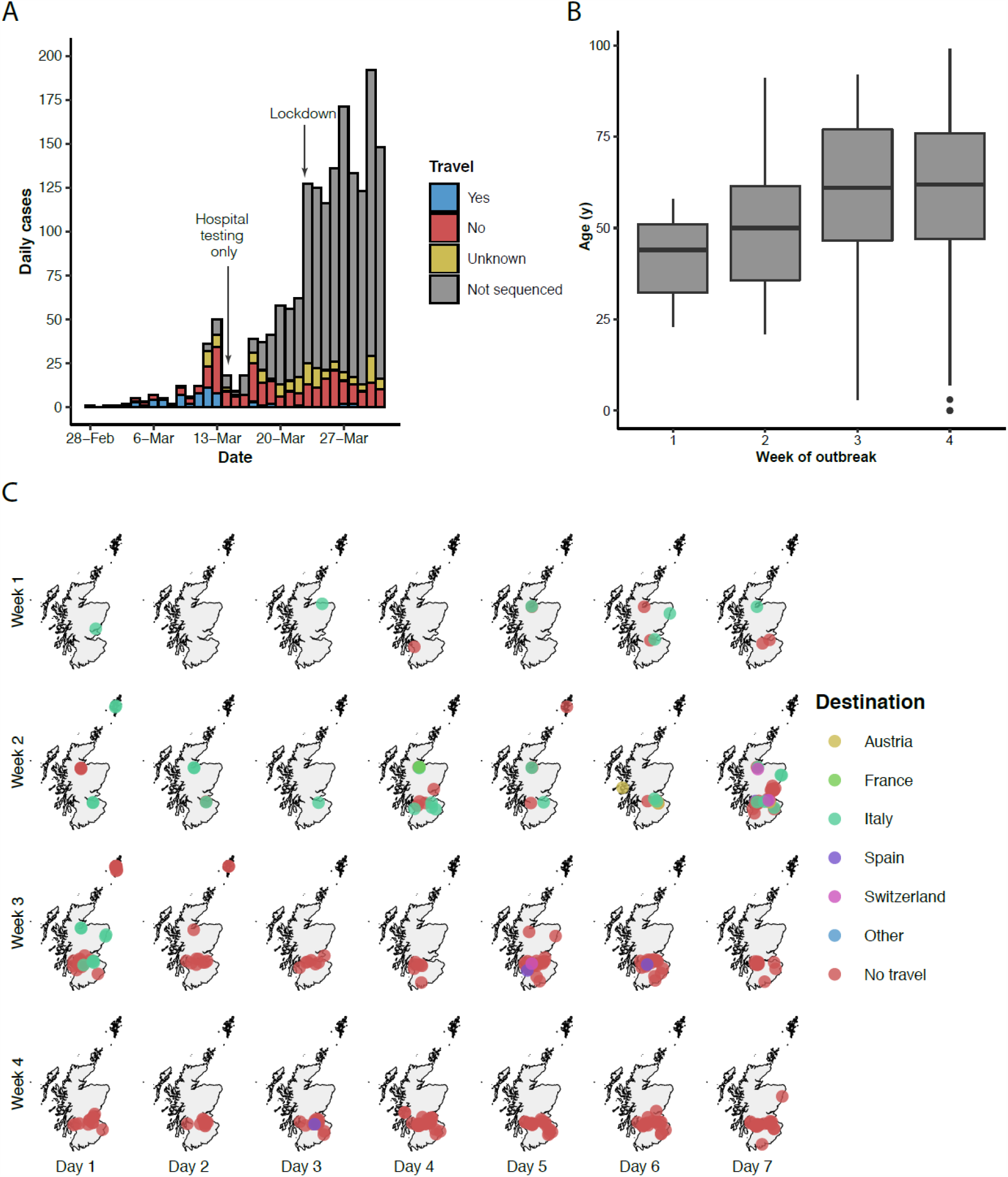
Spatial and demographic characteristics of cases that underwent SARS-CoV-2 sequencing in this study over the initial weeks of the Scottish outbreak. (A) Histogram of daily cases stratified by confirmed travel history. (B) Ages of positive cases by week of the outbreak in Scotland (C) Spatial distribution and associated travel history over the first four weeks of the outbreak generated using R. Testing of community cases ceased on day 1 of week 3 (14^th^ March) and lockdown occurred on day 2 of week 4 (23^rd^ March).

To determine the relationship of the viruses identified in Scotland to circulating SARS-CoV-2 variants, we inferred an evolutionary tree using viruses sequenced in this study against all complete genomes available from GISAID (Figure 2; Supplementary Fiigure 1). Overall limited variability in the genome was observed in keeping with the lower evolutionary rate of coronaviruses compared to other RNA viruses and the recent introduction of the virus into the human population. The 452 genomes displayed an average of 3.4 nonsynonymous and 1.8 synonymous nucleotide substitutions in comparison to Wuhan-Hu-1 (Figure 3). Analysis of sequencing error indicated that one genome out of seven re-sequenced using target enrichment contained a single insertion error, which would not have impacted clustering in the phylogenetic analysis. The strains introduced into Scotland are diverse with the majority fitting within the B lineage (n= 432) and only 20 from lineage A (22) (Figure 2). Two common amino acid replacements encoding 614D/G and 323P/L in spike and nsp12 respectively were observed. D614G has been hypothesised to be associated with increased transmissibility(24). Similarly another Spike replacement N439K (reported by the CoVsurver receptor binding surveillance GISAID, https://www.gisaid.org/covsurver) occurs with D614G and is circulating in a Scottish cluster of 12 sequences.

**Figure 2.**
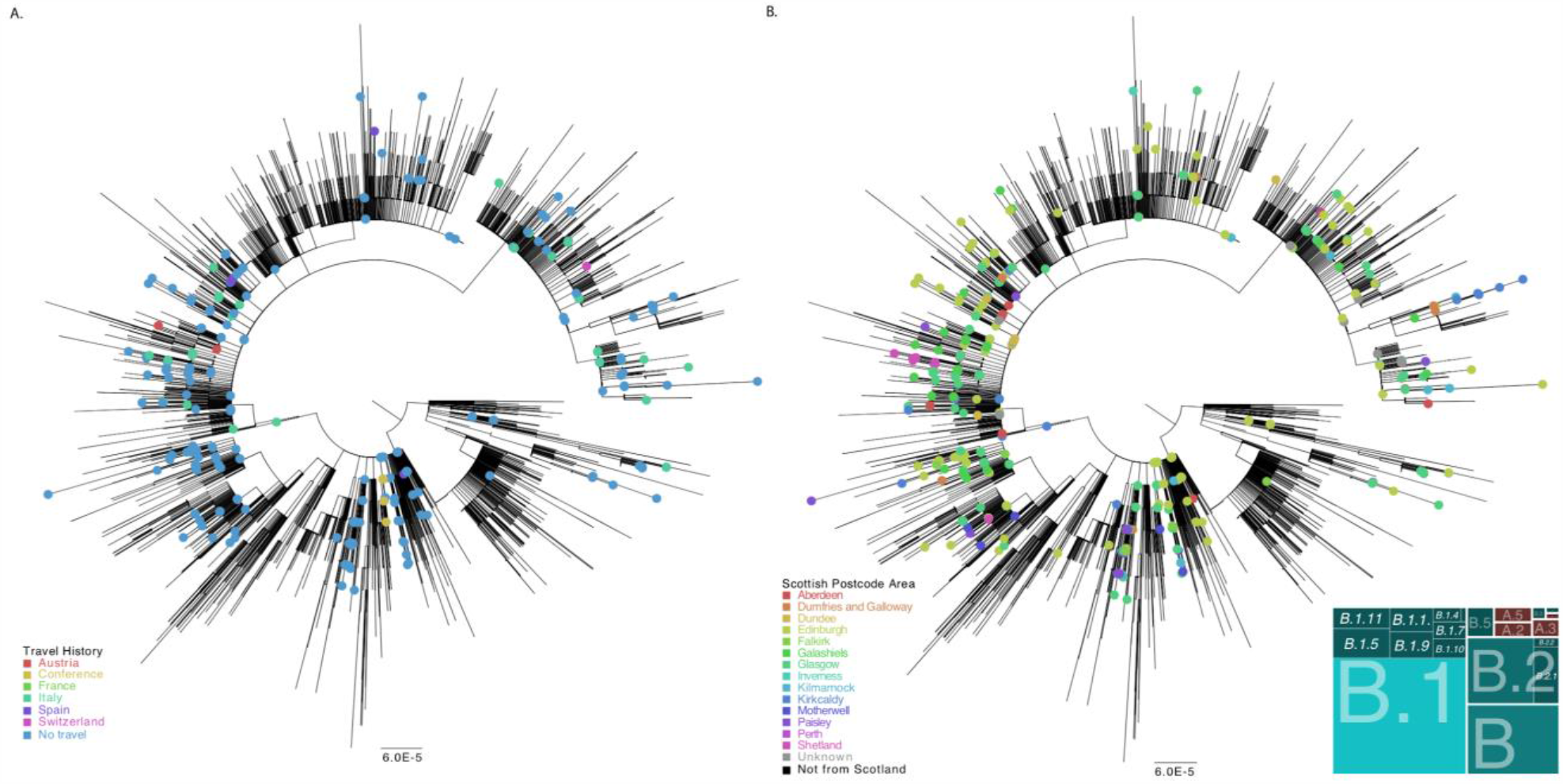
Phylogenetic trees showing the relationship of the 452 Scottish genomes to other SARS-CoV-2 genomes available from GISAID with known travel histories (A) and UK residence postal code areas (B) indicated (see keys). The panel in the bottom right indicates Scottish variant assignment within recently defined lineages. The scale bar indicates substitutions per nucleotide site.

**Figure 3.**
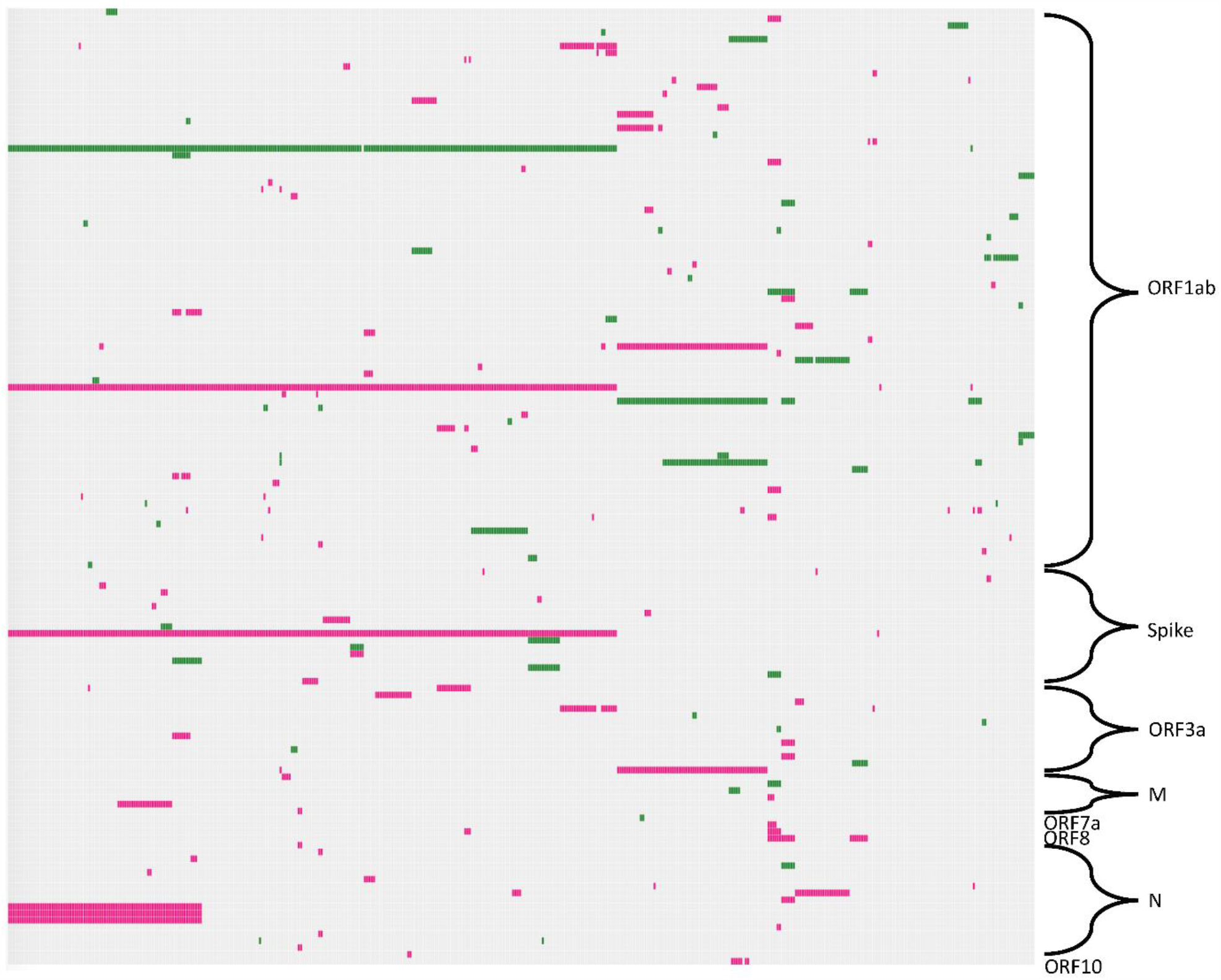
A visualisation of the genetic variation observed in the 452 SARS-CoV-2 genomes in Scotland. Nonsynonymous (pink) and synonymous (green) substitutions (with respect to Wuhan-Hu-1, GenBank accession number MN908947) are represented in colour in each row. The mutations are plotted in a grid format where each column is a sample and each row is a unique mutation at a given genome position; mutations have been filtered to only display those observed in more than one sample and labelled according to what ORF they are located in. The plot was created using the d3heatmap package in R, and the phylogram (top) displays the samples (columns) ordered according to Ward’s clustering.

Introductions were estimated based on both the phylogeny (22) and by travel history and date of sampling. The phylogenetic analysis combined with the epidemiological history permitted the identification of least 113 introductions into Scotland. Notably if identical basal sequences and singleton sequences without a significant travel history were to be counted as separate introductions, the phylogeny was suggestive of as many as 276 introductions into Scotland. Given some of the latter are probably linked infections the true number in our sample will be between these two numbers.

Many of the introductions of COVID-19 into Scotland were from known returning travellers from Europe, mostly Italy (37 out of 113 introductions). 38/86 (44%) of inferred introduction clusters were single cases not linked with further cases over time. Several singleton lineages were not associated with travel, likely corresponding to undetected community transmission clusters. 48 of the introductions resulted in clusters of at least two individuals and were associated with transmission in varied settings (Figure 4), for example, in the Shetland islands with travel links to Italy (Figure 4A), within a care home facility, community transmission across central Scotland and transmission related to an international conference event in Edinburgh at the end of February, prior to the first documented SARS-CoV-2 case in Scotland (Figure 5). The latter demonstrates the extent to which conferences and other large gatherings may contribute to the spread of the virus, supporting findings from similar events in China and Singapore (25, 26).

**Figure 4.**
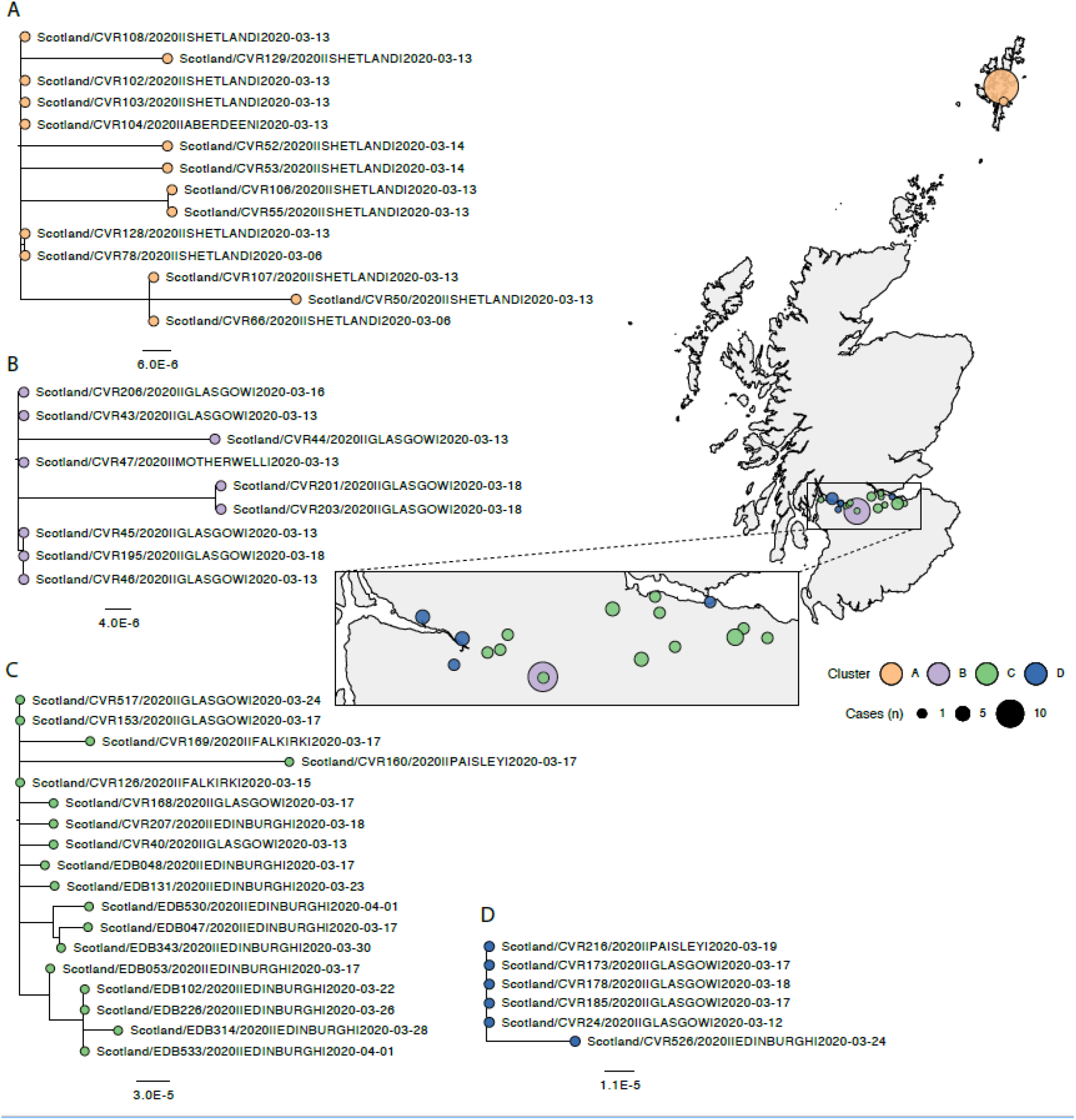
Examples of phylogenetic clusters associated with introduction events; (A) A cluster in Shetland associated with two linked cases with travel to Italy, (B) a focal outbreak in a residential facility not associated with any known travel, (C) a spatially distributed outbreak across the Central Scotland belt without any known links to travel, (D) a cluster associated with travel to Spain (D).

**Figure 5.**
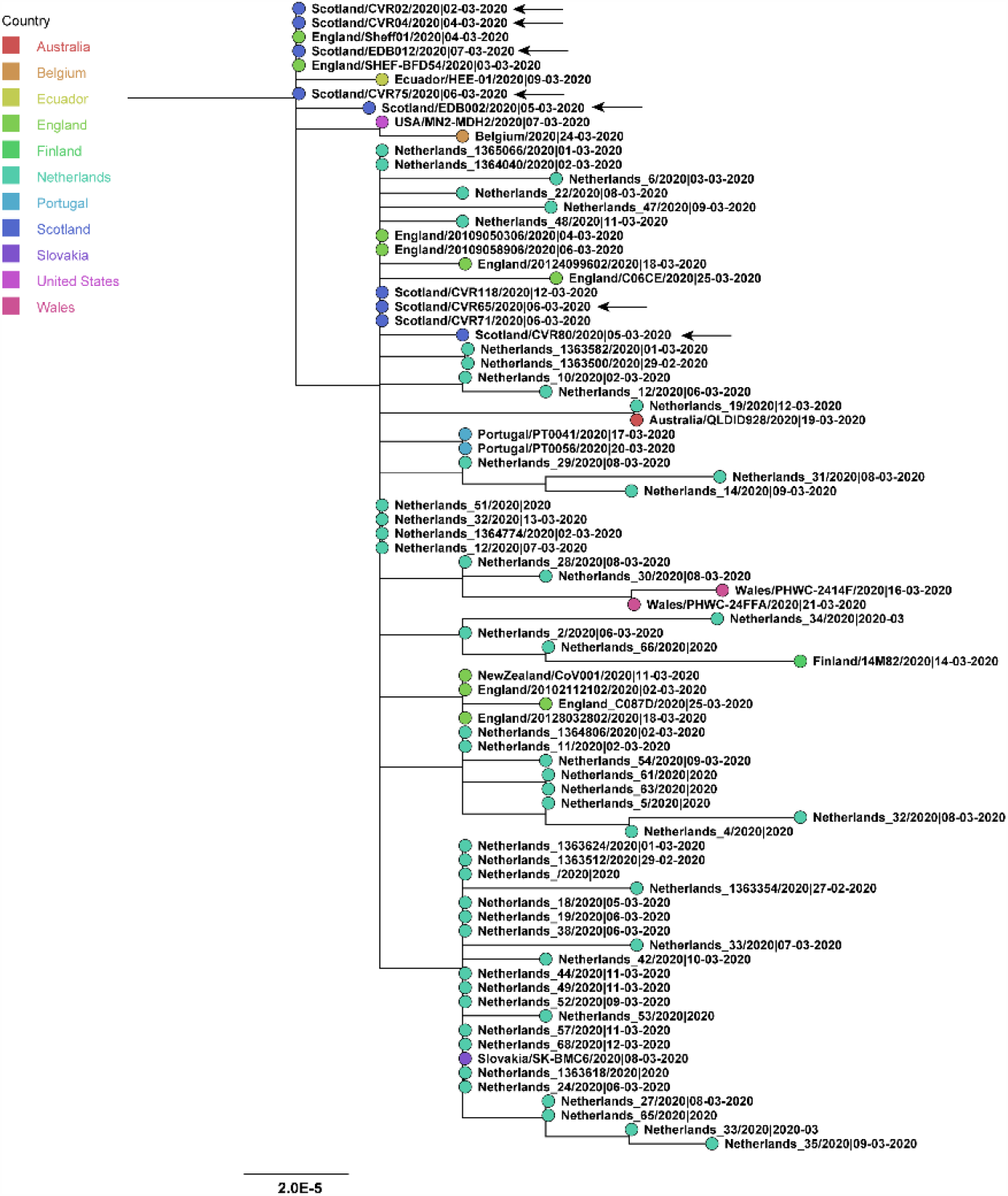
A phylogenetic cluster associated with an international event hosted in Scotland in February 2020 associated with international dispersal of this sub-lineage. Confirmed attendees are indicated by black arrows.

### Shift to community transmission

While early cases were most often associated with travel, later cases were almost all associated with community transmission in Scotland. Of 113 introductions, 50 (44%) had no associated travel links, 58 (51%) were linked to travel in Europe, three (3%) were linked to a Caribbean cruise and one was related to travel to Thailand and Italy. Of 86 phylogenetic clusters, 44 (51%) were linked to sequences from within the UK, outside Scotland. The first case of documented community transmission occurred on the 2^nd^ March 2020 and community transmission was well-established by the 11^th^ March (Figures 1, 4). A cluster occurring between the 12^th^ and 24^th^ March from Greater Glasgow contained an index case with travel history to Spain (Figure 4D) but all subsequent cases in the cluster did not report travel. This shift to community transmission is evident in other example clusters: Figure 4B represents a cluster from a care home, Figure 4C represents a large cluster which spread across the central belt between the 13^th^ March and 1^st^ April 2020 with no known associated travel.

We also investigated the possibility of local transmission chains in healthcare settings. A healthcare worker (HCW) working on a COVID-19 ward became infected at the beginning of the outbreak. To investigate the possibility of nosocomial transmission, all sequences from patients on the ward were compared with that of the HCW. However, the HCW, CVR10, showed evidence of infection from a virus strain distinct from other samples from the hospital within lineage B.2.2, whilst the ward patient sequences were from lineages B.1.5(CVR76), B.2.1(CVR07) and B.1.10(CVR79), indicating community rather than nosocomial infection (Supplementary Fiigure 2). While the phylogenetic signal is low owing to the low levels of variation in SARS-CoV-2 genome, this analysis demonstrates it is possible to exclude transmission events with high certainty. Such information is useful in complex settings such as health and care facilities where a case might indicate infection acquired elsewhere or a breakdown in control measures.

### Interpretation

Our study indicates that SARS-CoV-2 entered the Scottish population through at least 113 separate travel-related introductions, leading to at least 48 clusters of sustained community transmission and 38 singleton sequences with no evidence of onward transmission. 44/86 (49%) of phylogenetic clusters included sequences from individuals with a known history of international travel. The majority returned to Scotland from Europe at the end of February and early March following travel to Italy and less commonly to Spain, Austria, Switzerland, France, England, Ireland, Poland, the Caribbean and Thailand.

Travel to continental Europe in February and March 2020, the epicentre of the global COVID-19 pandemic, was a clear driver of the Scottish outbreak; the majority of the lineages detected in this study were related to European sequences. Cases with direct links to South-East Asia were rare. 20 lineage A sequences, a lineage present in China at the beginning of the outbreak (but which is now globally distributed) appear to have been introduced to Scotland via Spain. On the 28th January, the UK government recommended against all but essential travel to China and for returning travellers to self-isolate for two weeks upon their return regardless of symptoms (27). However, despite evidence of local transmission in Italy as early as February 21st, advice from the Scottish Government for returning travellers from Italy to self-isolate was issued only on the 25^th^ of February and was limited to those having returned from specific lockdown areas (28-30). By the time this advice was extended to all travellers on 10^th^ March, the COVID-19 outbreak within Scotland was already being driven by community transmission. A lack of robust measures to manage ingress of infected travellers from emerging pandemic hotspots may have accelerated the course of the outbreak in Scotland and the UK as a whole.

Our data suggest that SARS-CoV-2 was introduced to Scotland on multiple occasions, including at an international conference held in Edinburgh several days before the first Scottish case was confirmed. Whilst multiple local cases are linked to this event, the last case within the cluster in the UK occurred on the 27^th^ March, suggesting that the local public health response to this event may have been effective and did not account for the majority of cases in Scotland. However, the geographical distribution of other sequences is striking, spanning four continents and ten countries. The role of this event in dispersing the virus locally and internationally, before a single case had been identified in Scotland, demonstrates that governments should be wary of prematurely relaxing restrictions on large gatherings and international travel.

Importantly, we identified viral lineages with no epidemiological link to travel as early as two days after the first detection of infection, indicating earlier introduction to Scotland than the first detected case, reported on the 1^st^ March 2020 (23). 42/86 (49%) clusters were linked to sequences within the UK but outside Scotland. The majority of individuals had no recorded link to travel by the 12^th^ March, only 2 days after substantial travel and physical distancing restrictions were put in place. Importantly these data are suggestive of introductions and community spread before initial detection. The epidemic in the UK expanded rapidly, prompting the government to respond with restrictive public health measures or “lockdown” to disrupt transmission. As the number of cases subside, this phylogenetic data will provide a baseline for granular real-time sequencing of infections. This will be used as a measure of the success of current measures and has the potential to contribute to decision-making around the easing (or tightening) of public health measures.

Integration of genomic sequence data with traditional case-finding and contact tracing has the potential to enhance descriptive epidemiology and deliver more targeted control measures. However previous interventions of this type have been largely retrospective. The challenge will be to develop a framework where phylogenetic information is delivered in real-time in an easily actionable format to public health and infection control teams.

This study has some limitations. We sampled only 20% of those detected during the initial outbreak, therefore some introduction events will have been missed. Further, public health records were not accessed to record linkage with sequence data. While introduction of erroneous variation during sequencing or genome assembly is possible, we estimated experimentally that such rates were extremely low and should not affect clustering patterns in phylogenetic analyses. This analysis, based on the available phylogeny, is most likely to be an under-estimate, despite dense sampling of the beginning of the outbreak.

Linkage of sequences do not categorically prove that transmission events have occurred and require correlation with epidemiological information. Many events may be linked to an identical sequence because SARS-CoV-2 is a slowly evolving RNA virus. Exclusion of linkage may be inferred with more certainty. As the variation present in the global diversity is well-represented in Scotland, reflecting a high number of near simultaneous introductions, tracking of the outbreak is feasible and can be used to refine public health interventions. Our analysis may also have been affected by a shift in sampling all symptomatic individuals to hospitalised patients only during mid-March. Some travel-related introductions may therefore not have been detected (as evidenced by several clusters with no evidence of travel related infection after this time).

The utility of genomic epidemiology to inform local level policy is highlighted by the case of a healthcare worker in this study who tested positive for COVID-19 whilst working on a high-risk ward. The sequence from the healthcare worker was clearly distinct from those from the patients on the ward (Supplementary figure 2), suggesting that nosocomial transmission was unlikely. Such information can inform targeted infection control interventions such as the effectiveness of personal protective equipment (PPE). In contrast, several healthcare associated clusters were detected later in the outbreak that included healthcare workers.

In summary, the first month of the COVID-19 outbreak in Scotland was associated with multiple introductions related to returning travellers from Europe, increased evidence of ongoing non-travel related transmissions as time passed, and clusters related to events and healthcare facilities. Since the onset of lockdown interventions, introductions into the community have subsided. An earlier lockdown from countries with a high burden of cases such as Italy and other measures such as quarantine of travellers from high-risk areas might have prevented escalation of the outbreak and multiple clusters of ongoing community transmission.

## Data Availability

The data for the enclosed manuscript was made available in anonymised format to all authors.

## Acknowledgments

We would like to thank all Scottish virology laboratories who provided samples for sequencing, the authors who have deposited and shared genome data on GISAID, https://www.gisaid.org. Our genome sequence acknowledgments can be found in supplementary table 1. Edinburgh also thanks Richard Kuo, Tim Regan and Amanda Warr (Roslin Institute, University of Edinburgh) for the loan of sequencing reagents. Glasgow also thanks Scott Arkison for HPC maintenance.

## Supplementary Material

**Supplementary Figure 1**. Phylogenetic tree showing the relationship of the 452 Scottish genomes to other SARS-CoV-2 genomes with UK residence postal code areas indicated. The scale bar indicates substitutions per nucleotide site.

**Supplementary Figure 2**. Phylogenetic trees showing the placing of a health care worker (CVR10) relative to suspected transmitted cases in a hospital setting CVR07, CVR76 and CVR79. The scale bar indicates substitutions per nucleotide site.

**Supplementary Table 1**. GISAID SARS-CoV-2 genome sequence acknowledgements table.

**Supplementary Table 2**. Phylogenetic and epidemiological data analysis table.

## References

1. Committee WCH. Wuhan Municipal Health and Health Commission’s briefing on the current pneumonia epidemic situation in our city 2019. 2019.

2. Wu F, Zhao S, Yu B, Chen YM, Wang W, Song ZG, et al. A new coronavirus associated with human respiratory disease in China. Nature. 2020;579(7798):265–9.

3. Zhu N, Zhang D, Wang W, Li X, Yang B, Song J, et al. A Novel Coronavirus from Patients with Pneumonia in China, 2019. N Engl J Med. 2020.

4. Zhou P, Yang X-L, Wang X-G, Hu B, Zhang L, Zhang W, et al. A pneumonia outbreak associated with a new coronavirus of probable bat origin. Nature. 2020.

5. Gorbalenya AE, Baker SC, Baric RS, de Groot RJ, Drosten C, Gulyaeva AA, et al. The species Severe acute respiratory syndrome-related coronavirus: classifying 2019-nCoV and naming it SARS-CoV-2. Nature Microbiology. 2020.

6. Huang C, Wang Y, Li X, Ren L, Zhao J, Hu Y, et al. Clinical features of patients infected with 2019 novel coronavirus in Wuhan, China. Lancet. 2020.

7. Mao L, Jin H, Wang M, Hu Y, Chen S, He Q, et al. Neurologic Manifestations of Hospitalized Patients With Coronavirus Disease 2019 in Wuhan, China. JAMA Neurology. 2020.

8. Verdoni L, Mazza A, Gervasoni A, Martelli L, Ruggeri M, Ciuffreda M, et al. An outbreak of severe Kawasaki-like disease at the Italian epicentre of the SARS-CoV-2 epidemic: an observational cohort study. The Lancet. 2020.

9. WHO. Statement on the second meeting of the International Health Regulations (2005) Emergency Committee regarding the outbreak of novel coronavirus (2019-nCoV). World Health Organisation; 30 January 2020.

10. WHO. WHO Director-General’s opening remarks at the media briefing on COVID-19 - 11 March 2020. World Health Organisation; 11 March 2020.

11. Quick J, Loman NJ, Duraffour S, Simpson JT, Severi E, Cowley L, et al. Real-time, portable genome sequencing for Ebola surveillance. Nature. 2016;530(7589):228–32.

12. Grubaugh ND, Ladner JT, Lemey P, Pybus OG, Rambaut A, Holmes EC, et al. Tracking virus outbreaks in the twenty-first century. Nat Microbiol. 2019;4(1):10–9.

13. Kafetzopoulou LE, Pullan ST, Lemey P, Suchard MA, Ehichioya DU, Pahlmann M, et al. Metagenomic sequencing at the epicenter of the Nigeria 2018 Lassa fever outbreak. Science. 2019;363(6422):74–7.

14. Grubaugh ND, Ladner JT, Kraemer MUG, Dudas G, Tan AL, Gangavarapu K, et al. Genomic epidemiology reveals multiple introductions of Zika virus into the United States. Nature. 2017;546(7658):401–5.

15. Biek R, Pybus OG, Lloyd-Smith JO, Didelot X. Measurably evolving pathogens in the genomic era. Trends Ecol Evol. 2015;30(6):306–13.

16. Corman VM, Landt O, Kaiser M, Molenkamp R, Meijer A, Chu DK, et al. Detection of 2019 novel coronavirus (2019-nCoV) by real-time RT-PCR. Eurosurveillance. 2020;25(3):2000045.

17. Li H, Durbin R. Fast and accurate short read alignment with Burrows-Wheeler transform. Bioinformatics (Oxford, England). 2009;25(14):1754–60.

18. Grubaugh ND, Gangavarapu K, Quick J, Matteson NL, De Jesus JG, Main BJ, et al. An amplicon-based sequencing framework for accurately measuring intrahost virus diversity using PrimalSeq and iVar. Genome Biol. 2019;20(1):8-.

19. Katoh K, Misawa K, Kuma Ki, Miyata T. MAFFT: a novel method for rapid multiple sequence alignment based on fast Fourier transform. Nucleic Acids Research. 2002;30(14):3059–66.

20. Nguyen LT, Schmidt HA, von Haeseler A, Minh BQ. IQ-TREE: a fast and effective stochastic algorithm for estimating maximum-likelihood phylogenies. Mol Biol Evol. 2015;32(1):268–74.

21. Darriba D, Posada D, Kozlov AM, Stamatakis A, Morel B, Flouri T. ModelTest-NG: A New and Scalable Tool for the Selection of DNA and Protein Evolutionary Models. Molecular Biology and Evolution. 2019;37(1):291–4.

22. Rambaut A, Holmes EC, Hill V, O’Toole Á, McCrone J, Ruis C, et al. A dynamic nomenclature proposal for SARS-CoV-2 to assist genomic epidemiology. bioRxiv. 2020:2020.04.17.046086.

23. Hill KJ, Russell CD, Clifford S, Templeton K, Mackintosh CL, Koch O, et al. The index case of SARS-CoV-2 in Scotland. Journal of Infection. 2020.

24. Korber B, Fischer WM, Gnanakaran S, Yoon H, Theiler J, Abfalterer W, et al. Spike mutation pipeline reveals the emergence of a more transmissible form of SARS-CoV-2. bioRxiv. 2020:2020.04.29.069054.

25. Zhen-Dong T, An T, Ke-Feng L, Peng L, Hong-Ling W, Jing-Ping Y, et al. Potential Presymptomatic Transmission of SARS-CoV-2, Zhejiang Province, China, 2020. Emerging Infectious Disease journal. 2020;26(5):1052.

26. Pung R, Chiew CJ, Young BE, Chin S, Chen MIC, Clapham HE, et al. Investigation of three clusters of COVID-19 in Singapore: implications for surveillance and response measures. The Lancet. 2020;395(10229):1039–46.

27. FCO. Foreign & Commonwealth Office; 28 January 2020 [Available from: https://www.gov.uk/government/news/fco-advises-against-all-but-essential-travel-to-mainland-china.

28. Government S. Preparations for coronavirus stepped up: Scottish Government; 25 February 2020 [Available from: https://www.gov.scot/news/preparations-for-coronavirus-stepped-up/.

29. ECDC. Outbreak of novel coronavirus disease 2019 (COVID19): situation in Italy – 23 February 2020. Stockholm: European Centre for Disease Prevention and Control; 23 February 2020.

30. ECDC. Communicable disease threats report; 16-22 February 2020, week 8. Stockholm: European Centre for Disease Prevention and Control; 21 Feb 2020.

